# Evaluation of the efficacy of using indocyanine green associated with fluorescence in sentinel lymph node biopsy

**DOI:** 10.1101/2022.08.29.22278953

**Authors:** Rafael da Silva Sá, Raquel Fujinohara Von Ah Rodrigues, Luiz Antônio Bugalho, Suelen Umbelino da Silva, Afonso Celso Pinto Nazário

## Abstract

Introduction: Sentinel lymph node biopsy is the established technique in the axillary staging of patients with early breast cancer without clinical axillary involvement. Three techniques are widely used globally for the detection of sentinel lymph nodes: patent blue; the radiopharmaceutical technetium 99 with the use of the gamma probe; and the combination of these two techniques. Objectives: To evaluate the sentinel lymph node detection rate with an innovative technique: indocyanine green associated with fluorescence in breast cancer patients, its comparison with the other methods (patent blue and combined patent blue + indocyanine green). Methods: Ninety-nine patients were subdivided into three arms with 33 patients, one arm was submitted to the sentinel lymph node technique using patent blue, the other indocyanine green, and the third a combination of the two dyes. Results: The accuracy rate in the identification of the sentinel lymph node was 78.8% with the use of patent blue, 93.9% with indocyanine green and 100% with patent blue + indocyanine green. The combined group identified mainly two sentinel nodes (48.5%); however, the other groups more commonly identified only one sentinel lymph node. The mean time of sentinel lymph node identification was 20.6 minutes among patients submitted to the traditional dye, 8.6 minutes in the indocyanine green arm and 10 minutes in the combination of the two methods (P<0.001). The mean surgery time was 69.4 minutes with patent blue, 55.1 minutes with indocyanine green and 69.4 minutes with the combination (P<0.001). Conclusions: The sentinel lymph node detection rate by fluorescence with the use of indocyanine green was considered effective. The comparison of the sentinel lymph node detection rate between the use of patent blue, indocyanine green and patent blue + indocyanine green (combined) revealed statistically significant differences (p=0.030), with the combined method being the most effective.

## Introduction

The treatment of breast cancer is a challenge to global public health as it is the most diagnosed malignant neoplasm in the world, reaching an annual estimate of 2.26 million new cases. It is the leading cause of cancer mortality among women. Adding to this, and to the worldwide alarm: two thirds of the affected population reside in undeveloped countries^1^.

The surgical treatment of breast cancer includes, in most cases, axillary surgery. Lymph node research is not indicated in conservative surgeries in patients with ductal carcinoma *in situ*^2^ or may be omitted in selected senile patients^3^. Three techniques are widely used globally for the detection of sentinel lymph nodes: patent blue, the radiopharmaceutical technetium 99 with the use of the gamma probe, and the combination of these two techniques. Patent blue, published by Giuliano et al in 1994^4^, was applied between 0.5 and 10 ml in the peritumoral region with a 25-gauge needle soon after anesthetic induction. Initially, the detection rate was 65.5% (114 out of 174 cases)^4^, which was quickly improved by Kern et al in 1999 with rates above 98% (38 out of 40 cases)^5^. The identification of the sentinel lymph node using technetium 99-m(99mTc) associated with the gamma probe, described by Krag et al in 1993, was possible with the peritumoral application of this marker 1 to 9 hours before surgery at a dose of 0.4 mCi associated with 0.5 ml of saline solution. The gamma detector initially used was the C-Trak from Care Wise Medical Products (Morgan Hill, CA, USA)^6^. The detection rate of 89% was published by Borgstein et al in 1998^7^.

These two dyes are so present in the routine of breast surgeons, especially after the important NSABP B-32 study, which reiterated the safety of the sentinel lymph node technique, using one of these two dyes in all its 80 research centers^8^. Each of these techniques has its strengths and weaknesses. The main defect of patent blue is the dermal tattoo, which can be eternal. Technetium 99, in turn, is expensive and requires a nuclear medicine team. With the desire to publicize another more modern dye option, Kitai et al were the pioneers to report, in 2005, the technique known as ICG (Indocyanine Green) in breast cancer and obtained an excellent identification rate of 94%. ICG is a low molecular weight non-radioactive fluorescent dye. This chemical marker can penetrate human tissue to depths of a few millimeters to a few centimeters. This allows real-time lymphatic migration, helping the surgeon to plan the dermal incision, thus reducing the difficulty of the procedure ^9^.

ICG is a tricarbocyanine, two heterocyclic rings joined by 7 carbons, with the following molecular formula: C43H47N2NaO6S2^10^. An innovative procedure raises concerns about patient safety. Indocyanine green is contraindicated in patients allergic to iodine and its derivatives, due to the potential risk of anaphylaxis^11^.

In fact, ICG was developed in 1955 and approved for clinical use in 1959 in the United States, with an indication outside the oncological scenario. However, its use in ophthalmology for retinal angiography intensified in the 1970s. Its essence as a fluorescent marker consists of illuminating the tissue of interest at the excitation wavelength (750 to 800 nm), having filters to spectrally select the fluorescence before reaching the sensor, presenting greater emission around 800 nm and at longer wavelengths depending on concentration, pH and temperature. The use of ICG in breast surgery is still a little used procedure^12^. Some studies have already validated the ICG for the use of sentinel lymph nodes with results as good as or even better than radioisotopes. However, the main ones were published by Asian countries in Asia (China and Japan, for example, already use it on a large scale) and in Europe only a few countries developed studies in relatively small cohorts, and this innovative technique has not yet replaced the pioneers with the justification that there are no local studies enough to prove the non-inferiority of ICG in relation to patent blue and 99mTc^13^.

This research presents the prospect of disseminating the results of an innovative and still little known technique for sentinel lymph node research. The fact that the guidelines of the main oncology entities worldwide, ASCO^14^ (American Society of Clinical Society), ESMO^15^ (European Society of Clinical Oncology) and NCCN®^16^ (The National Comprehensive Cancer Network) still do not include this dye as an option in their protocols reinforces the importance of this study.

## Materials and Methods

### Study design, setting and ethics

This study is a non-randomized clinical trial (prospective study with non-blinded sequential allocation of cases) performed in patients with early breast cancer with clinically negative axilla who had an indication for sentinel lymph node investigation during breast surgery. Exclusion criteria were: presence of staging T4, N1, N2 and N3; having undergone neoadjuvant chemotherapy; and having a history of allergy to iodine and its derivatives. This research was developed by the Discipline of Mastology, Department of Gynecology, Escola Paulista de Medicina / Universidade Federal de São Paulo and carried out at Hospital de Esperança - Hospital Regional do Câncer de Presidente Prudente.

The free and informed consent form was duly signed by all patients submitted to the research, after a thorough explanation of the study and procedures to be performed. The researchers ensured the confidentiality of all patients, not publicly disclosing the name or any other information that could identify the women involved in this study.

This research was approved by the Research Ethics Committee of the Federal University of São Paulo (CAAE: 08169419.9.0000.5505) and Research Ethics Committee of Hospital de Esperança - Regional Cancer Hospital of Presidente Prudente (CAAE: 08169419.9.3001.8247).

### Materials used

The materials used were: patent blue dye v 50mg/2ml, brand Pharmédice®, indocyanine green dye 5ml/10ml, brand OPHTALMOS®, 5ml syringe, brand SR®, needle 40 × 12 (dye aspiration) and needle 30 × 8 (periareolar infiltration), both branded SR®. For fluorescence, an image1S® device coupled to a D-light P® light source and a laparoscopy camera, all from the German company Karlz-Storz®, were used.

### Data sources and procedures performed

Data were obtained by manually filling in the patient’s data collection form, previously prepared with the main parameters that would be analyzed by the study.

After the information was collected, all of these were inserted into a specific Microsoft Excel® “template”, prepared by the statistician.

Patients in the well-known patent blue technique group underwent periareolar infiltration of 2 ml of this product with subsequent five-minute breast massage for migration of the lymphotropic dye during anesthetic induction. After this period, during the axillary dissection, the lymphatic ducts were visualized up to the sentinel lymph node (first lymph node of the lymphatic drainage of the breast), which was resected.

Patients in the innovative indocyanine green technique group underwent periareolar infiltration of 10 ml/5mg of this product, followed by a five-minute breast massage for migration of the fluorescent dye, also immediately following anesthetic induction. After this period, in the axillary dissection, the lymphatic ducts up to the sentinel lymph node were visualized using a Karlz-Storz® fluorescence device called image 1S®, coupled to a D-light P® light source and a camera of real-time visualization that has infrared light with 760 nm, coupled to a laparoscopy fluorescence camera. There was no damage to image quality, since this device performs fluorescence in closed surgeries and in the literature, there is already a description of its use also in open breast surgeries^17^.

The patients in the group with the combined technique underwent the two associated procedures previously described.

The access route for the identification of the sentinel lymph node was the best from an aesthetic point of view. Thus, some patients had only one breast incision (superior lateral quadrant, for example) and others had two breast incisions. In this regard, we tried not to change the surgeons’ routine and preference.

### Patient selection

Patients were allocated by sequence to the three study arms between 8/2/2019 and 12/16/2021. The study was designed with the proposed sample of 99 patients, as it is the mean N of the main fluorescence studies in the breast cancer scenario ^9,17,18^. The first 33 patients underwent sentinel lymph node biopsy using the patent blue technique. Patient number 34 to patient number 66 were included in the indocyanine green arm and finally the other patients (67 to 99) participated in the group that received patent blue + indocyanine green. This was necessary, because during the beginning of the research, the surgical center used for all patients at the Regional Cancer Hospital of Presidente Prudente was the Santa Casa de Misericórdia de Presidente Prudente, an attached hospital and maintainer of the oncology hospital. The surgical center of the Regional Cancer Hospital of Presidente Prudente, which includes fluorescence technology, was inaugurated in October/2019, a few months after the start of the research and the full release of its use as a routine occurred only in 2020. Therefore, in order to maintain the sequential allocation, the second arm was the indocyanine green arm, thus the third and final arm was the combined arm. We believe that this fact did not affect the accuracy of the study, as the patients were admitted upon arrival at the Mastology service.

### Data collection

We analyzed at the time of surgery:

- Time of identification of the sentinel lymph node (the first dermal incision was considered as the time of onset). We used a digital clock on the wall of the operating room to measure the time.
- Number of lymph nodes identified.
- Total surgery time (procedure time was considered from the first dermal incision to the end of the last dermal suture). The same digital clock in the surgical procedure room was used to accurately measure time.
- Intraoperative complications.

We analyzed on the 7th postoperative day:

- Measurements of the scar(s).
- Portovac EZ-Suc drain flow from the Cremer® brand.

We analyzed the presence or absence of dermal tattoo on the 60th postoperative day and adverse events at follow-up.

Axillary status was defined according to the number of positive lymph nodes.

The cutoff value for the Ki-67 was 14% score. Above this value it was considered as high Ki-67 and below 14% as low.

Tumor grade was classified using the modified Bloom-Richardson system^19^ and staging was performed using the AJCC Cancer Staging Manual 8th Edition^20^.

The economic impact of the procedure was evaluated through a thorough financial analysis of the costs that Hospital de Esperança had for each patient submitted to the research. In this survey by the financial department, all the amounts paid during the entire hospitalization were analyzed: fees, food, medication, serum therapy, pathology, breast implants and the dyes used.

### Statistical analysis

Data were presented in terms of their frequencies and percentages for categorical variables, mean and standard deviation for continuous variables and median (minimum – maximum) for discrete variables. In the comparisons of the variables between the groups of dyes, the ANOVA test was used for normal variables, followed by the Tukey test as posthoc; Kruskal-Walis test for non-normal; and Chi-Square test (or Fisher’s exact test when appropriate) for the categorical ones. The significance level adopted in all tests was 5%. To assist in the analysis, the R software, version 4.1.3 was used.

To compare the values spent according to the type of dye, the ANOVA test and the Tukey test were used for multiple comparisons.

## Results

The total N proposed for the study of 99 patients was met, with 3 arms with 33 patients each. The mean age was 58.4 years (standard deviation of 13.3). The most affected quadrant was the right superior lateral in 21.2% (n=21), followed by the left superior lateral in 14.1% (n=14). During the study period, there were no reports of male patients or patients with ductal carcinoma *in situ*. The vast majority of patients, 70.7% (n=70), were already post-menopausal and the remainder (29.3%, n=29) were still in menacme (S1 Table).

We also evaluated the BMI (body mass index) of the patients, believing in a possible interference in this index with the dose/type of dye used. Most patients were over-weight during the evaluation of the first medical appointment in 44.4% (n=44).

Three surgical techniques were performed: breast-conserving surgery in 84.8% (n=84), mastectomy without breast reconstruction in 10.1% (n=10) and mastectomy with breast reconstruction (expander) in 5.1% (n=5).

The histological subtypes had the following distribution: non-special invasive carcinoma 92.9% (n=92), invasive lobular carcinoma 4% (n=4), invasive papillary carcinoma 2% (n=2) and invasive medullary carcinoma 1% (only 1 case). The tumor size was classified between T1b, T1c and T2, being represented in the sample mainly the T1c, in 53.5% (n=53). No cases of T1a and T3 were reported. The immunohistochemical profile was classified into: Luminal B 35.4% (n=35), Luminal A 37.4% (n=37), Luminal HER 12.1% (n=12), Triple Negative 8.1% (n=8) and HER 2+ 7.1% (n=7).

The number of lymph nodes identified was mainly one in 46.9% of cases (n=46) and two in 35.7% (n=35). The median was one lymph node (0-4). Nine cases in total had no lymph nodes identified by the techniques used. The median of resected lymph nodes was three. The median of positive lymph nodes was zero (Table 1).

**Table 1.** Surgical aspects of the three sentinel lymph node detection methods (patent blue, indocyanine green and combined).

| Characteristic | n = 99 (%) |
| --- | --- |
| **Dye** |  |
| Patent blue | 33 (33.3%) |
| Indocyanine green | 33 (33.3%) |
| Patent blue + indocyanine green | 33 (33.3%) |
| **Number of lymph nodes identified** |  |
| 0 | 9 (9.2%) |
| 1 | 46 (46.9%) |
| 2 | 35 (35.7%) |
| 3 | 7 (7.1%) |
| 4 | 1 (1.0%) |
| Median (min - max) | 1 (0 - 4) |
| **Number of lymph nodes resected** |  |
| Median (min - max) | 3 (0 - 27) |
| **Number of lymph nodes +** |  |
| Median (min - max) | 0 (0 - 17) |
| **Identification time (min)** |  |
| Mean ± SD | 12.7 ± 10.1 |
| **Surgery time (min)** |  |
| Mean ± SD | 64.6 ± 18.0 |
| **Intraoperative complications** |  |
| Dye allergy | 2 (2.0%) |
| **Number of incisions** |  |
| 1 | 55 (55.6%) |
| 2 | 44 (44.4%) |
| **Breast incision measurement (cm)** |  |
| Mean ± SD | 6.1 ± 1.4 |
| **Axillary incision measurement (cm)** |  |
| Mean ± SD | 5.5 ± 8.8 |
| **Single breast incision measurement (cm)** |  |
| Mean ± SD | 8.3 ± 3.9 |
| **Drain output in 7 days (ml)** |  |
| Mean ± SD | 485.1 ± 337.5 |
| **Adverse events at follow-up** |  |
| Surgical wound dehiscence | 5 (8.3%) |
| Radio-induced scleroderma | 1 (1.7%) |
| Hematoma | 4 (6.7%) |
| Infection + Lung Metastasis | 1 (1.7%) |
| Local Infection + Dermal Tattoo | 1 (1.7%) |
| Bone metastasis | 2 (3.3%) |
| Bone + liver metastasis | 1 (1.7%) |
| Liver + lung metastasis | 1 (1.7%) |
| Relapse | 1 (1.7%) |
| Relapse + Bone metastasis | 1 (1.7%) |
| Relapse + Bone + liver + brain metastasis | 1 (1.7%) |
| Seroma | 6 (10%) |
| Seroma + Local Infection | 1 (1.7%) |
| Tattoo on breast skin | 34 (56.7%) |

Adding all the arms together, the mean time for sentinel lymph node identification was 12.7 minutes and the total surgery time was 64.6 minutes.

55.6% of patients received only 1 breast incision. The other patients underwent 2 dermal incisions. The mean size of the breast incision was 8.3 cm when the incision was single. When 2 incisions were performed, the average measurement of the breast incision was 6.1 cm, and the axillary incision was 5.5 cm.

The average output of the Portovac drain in 7 days was 485.1ml. There were no intraoperative complications and the main adverse event in the follow-up was dermal tattoo in 34 cases, corresponding to 56.7% of the patients.

Comparison between three arms did not reveal a statistically significant difference between the following characteristics (S2 Table):

- Age (P=0.717).
- Menopause status (P=0.220).
- BMI (P=0.875).
- Type of surgery (P=0.555).
- Anatomical staging (P=0.063).
- Prognostic staging (P=0.090).
- Histological subtype (P=0.346).
- Histological grade (P=0.786).
- Immunohistochemical analysis (P=0.377).

The comparison between the three different dye techniques identified a statistically significant difference in relation to the affected quadrant (P=0.026), with the right superior lateral quadrant being the most present in the patent blue group (33.3%), the left superior lateral quadrant in the indocyanine green group (21.2%) and the right superior lateral quadrant together with the union of the upper quadrants of the left breast (15.2% each) in the combined arm. Tumor size also showed differences (P=0.046), although T1c was the main stage classified in all groups.

The accuracy rate in identifying the sentinel lymph node was 78.8% with the use of patent blue, 93.9% with indocyanine green and 100% with patent blue + indocyanine green (Fig 1). The combined group identified mainly 2 sentinel nodes (48.5%), however the other groups more commonly identified 1 sentinel node only (Table 2).

**Table 2.**
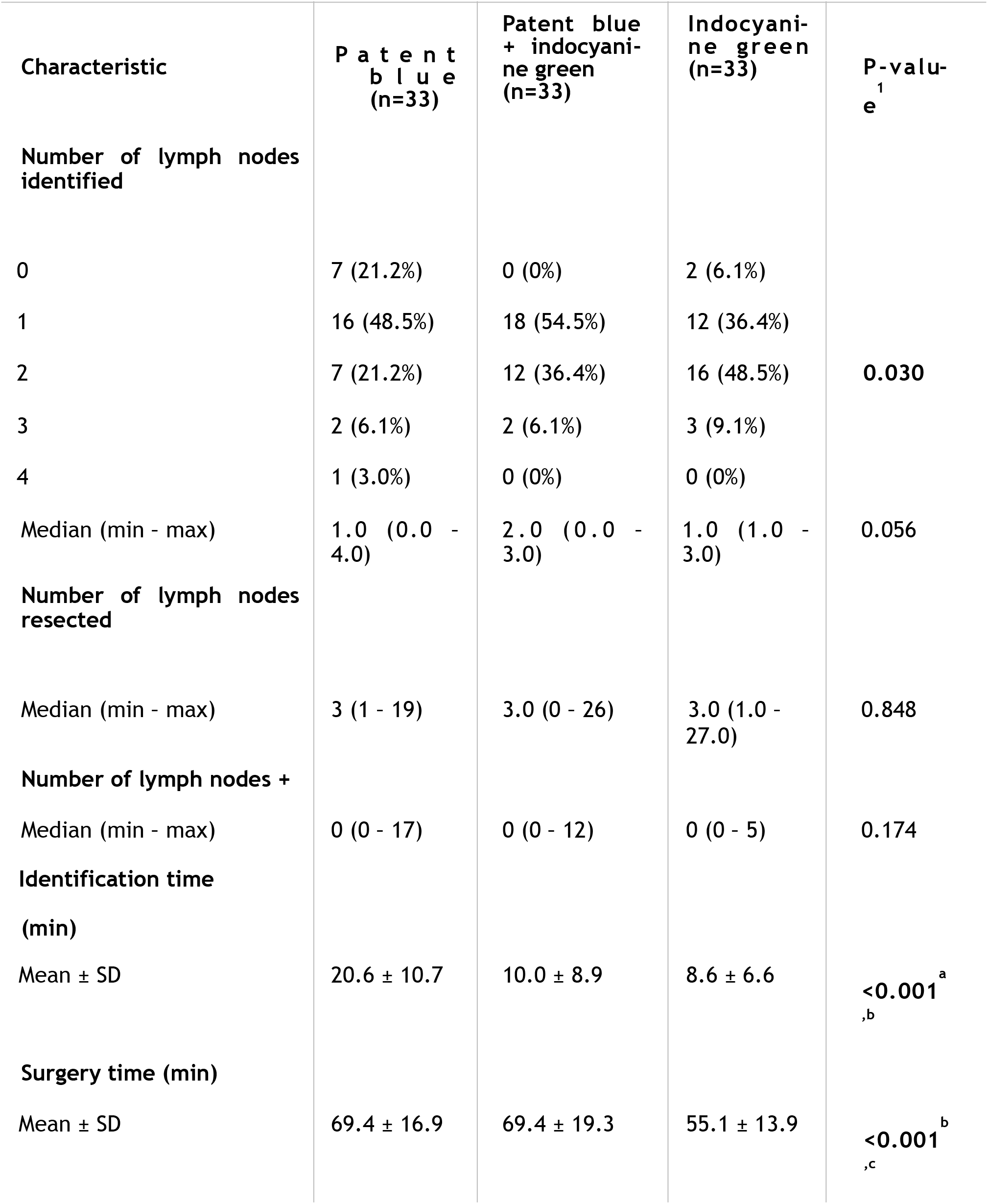
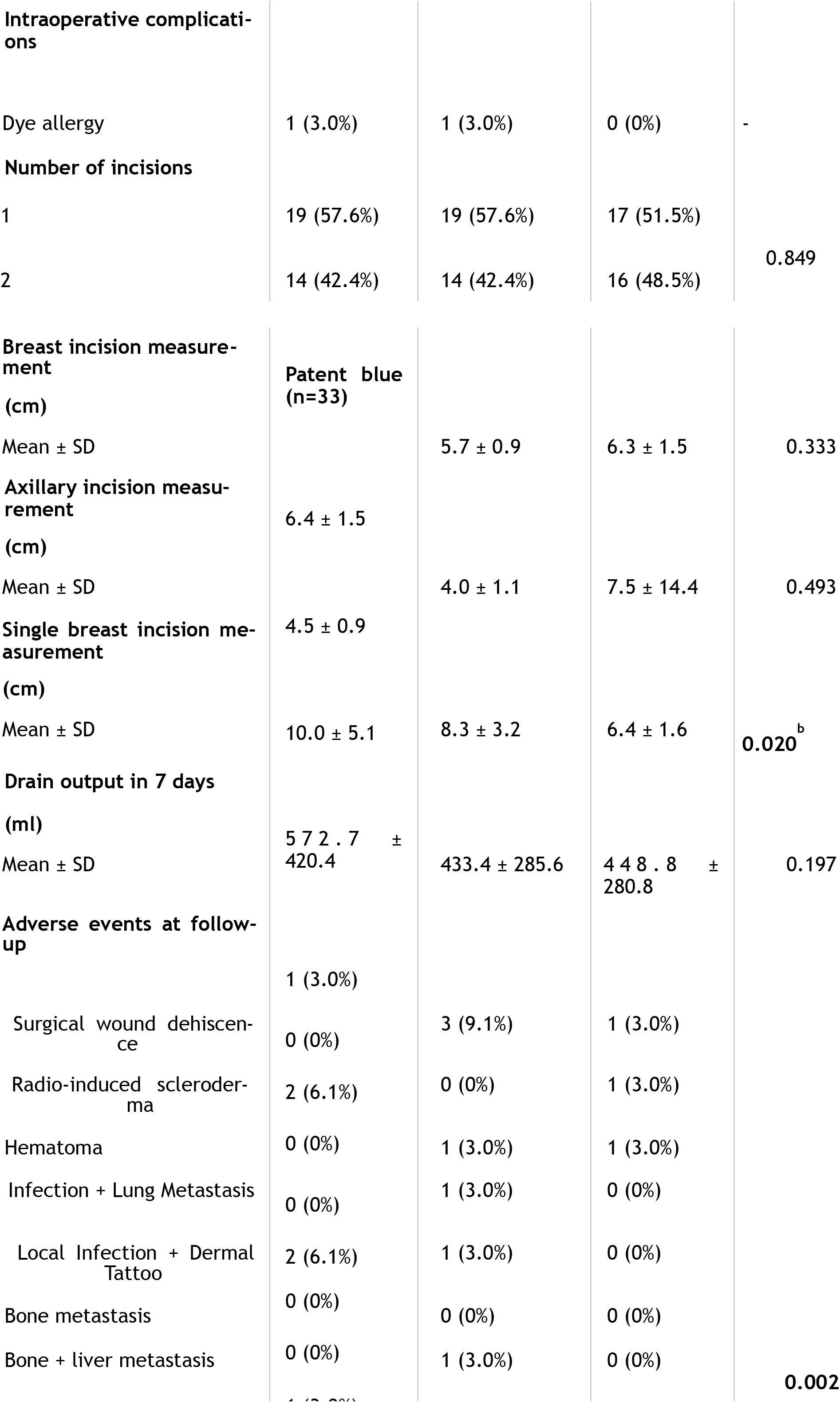
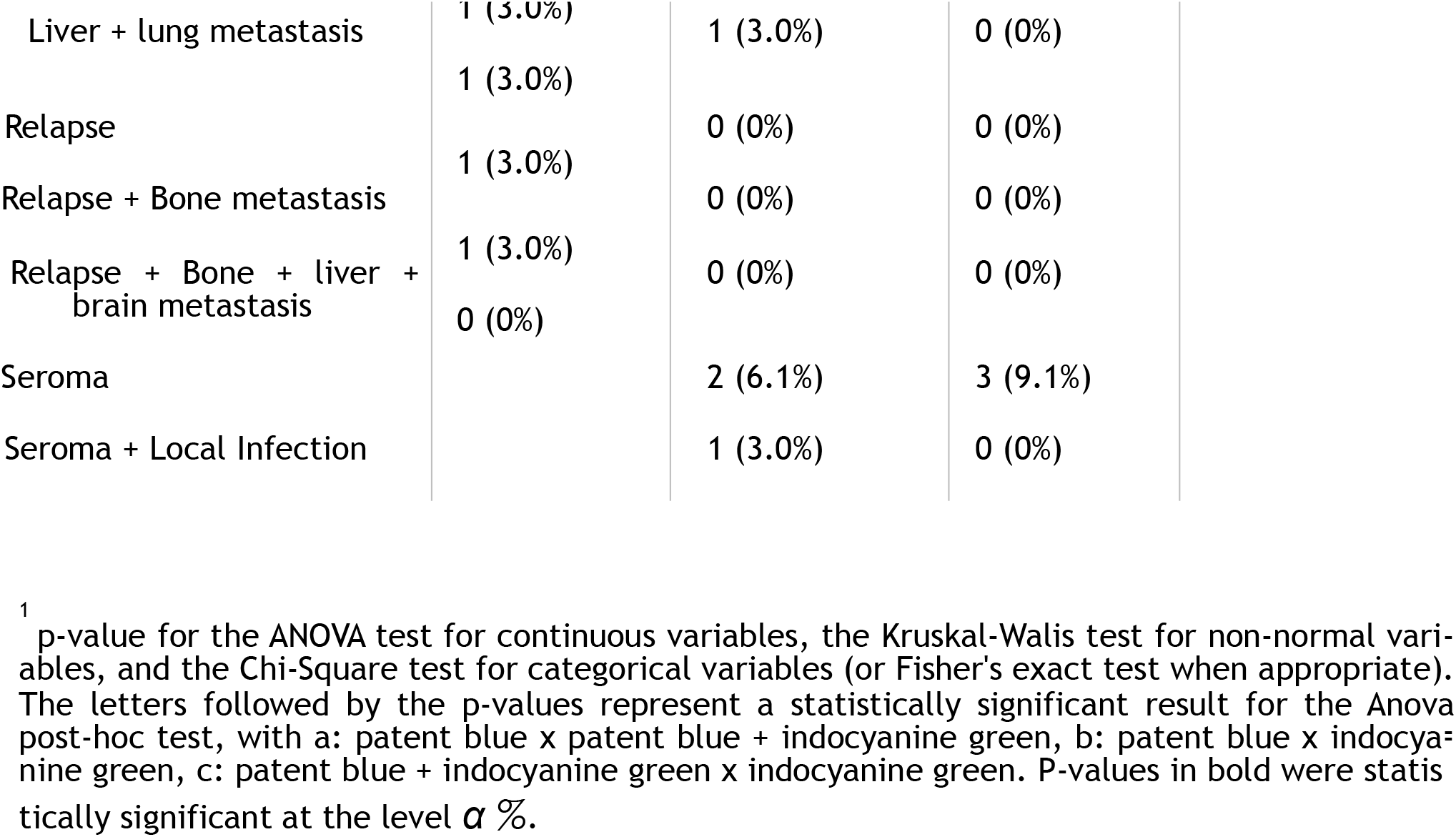
Comparison of surgical aspects of the three sentinel lymph node detection methods (patent blue, indocyanine green and combined).

| Characteristic | P a t e n t<br>b l u e<br>(n=33) | Patent blue<br>+ indocyanine<br>green<br>(n=33) | Indocyanine<br>green<br>(n=33) | P-value<sup>1</sup><br>e |
| --- | --- | --- | --- | --- |
| **Number of lymph nodes identified** |  |  |  |  |
| 0 | 7 (21.2%) | 0 (0%) | 2 (6.1%) |  |
| 1 | 16 (48.5%) | 18 (54.5%) | 12 (36.4%) |  |
| 2 | 7 (21.2%) | 12 (36.4%) | 16 (48.5%) | **0.030** |
| 3 | 2 (6.1%) | 2 (6.1%) | 3 (9.1%) |  |
| 4 | 1 (3.0%) | 0 (0%) | 0 (0%) |  |
| Median (min - max) | 1.0 (0.0 - 4.0) | 2.0 (0.0 - 3.0) | 1.0 (1.0 - 3.0) | 0.056 |
| **Number of lymph nodes resected** |  |  |  |  |
| Median (min - max) | 3 (1 - 19) | 3.0 (0 - 26) | 3.0 (1.0 - 27.0) | 0.848 |
| **Number of lymph nodes +** |  |  |  |  |
| Median (min - max) | 0 (0 - 17) | 0 (0 - 12) | 0 (0 - 5) | 0.174 |
| **Identification time (min)** |  |  |  |  |
| Mean ± SD | 20.6 ± 10.7 | 10.0 ± 8.9 | 8.6 ± 6.6 | **<0.001<sup>a</sup>**<br>,b |
| **Surgery time (min)** |  |  |  |  |
| Mean ± SD | 69.4 ± 16.9 | 69.4 ± 19.3 | 55.1 ± 13.9 | **<0.001<sup>b</sup>**<br>,c |
| **Intraoperative complications** |  |  |  |  |
| Dye allergy | 1 (3.0%) | 1 (3.0%) | 0 (0%) | - |
| **Number of incisions** |  |  |  |  |
| 1 | 19 (57.6%) | 19 (57.6%) | 17 (51.5%) | 0.849 |
| 2 | 14 (42.4%) | 14 (42.4%) | 16 (48.5%) |  |
| **Breast incision measurement (cm)** | **Patent blue (n=33)** |  |  |  |
| Mean ± SD | | 5.7 ± 0.9 | 6.3 ± 1.5 | 0.333 |
| **Axillary incision measurement (cm)** | 6.4 ± 1.5 | | | |
| Mean ± SD | | 4.0 ± 1.1 | 7.5 ± 14.4 | 0.493 |
| **Single breast incision measurement (cm)** | 4.5 ± 0.9 | | | |
| Mean ± SD | 10.0 ± 5.1 | 8.3 ± 3.2 | 6.4 ± 1.6 | 0.020<sup>b</sup> |
| **Drain output in 7 days (ml)** |  |  |  |  |
| Mean ± SD | 572.7 ± 420.4 | 433.4 ± 285.6 | 448.8 ± 280.8 | 0.197 |
| **Adverse events at follow-up** |  |  |  |  |
|  | 1 (3.0%) |  |  |  |
| Surgical wound dehiscence | 0 (0%) | 3 (9.1%) | 1 (3.0%) |  |
| Radio-induced scleroderma | 2 (6.1%) | 0 (0%) | 1 (3.0%) |  |
| Hematoma | 0 (0%) | 1 (3.0%) | 1 (3.0%) |  |
| Infection + Lung Metastasis | 0 (0%) | 1 (3.0%) | 0 (0%) |  |
| Local Infection + Dermal Tattoo | 2 (6.1%) | 1 (3.0%) | 0 (0%) |  |
| Bone metastasis | 0 (0%) | 0 (0%) | 0 (0%) |  |
| Bone + liver metastasis | 0 (0%) | 1 (3.0%) | 0 (0%) | 0.002 |

|  |  |  |  |
| --- | --- | --- | --- |
| Liver + lung metastasis | 1 (3.0%) | 1 (3.0%) | 0 (0%) |
|  | 1 (3.0%) |  |  |
| Relapse |  | 0 (0%) | 0 (0%) |
|  | 1 (3.0%) |  |  |
| Relapse + Bone metastasis |  | 0 (0%) | 0 (0%) |
|  | 1 (3.0%) |  |  |
| Relapse + Bone + liver + brain metastasis |  | 0 (0%) | 0 (0%) |
|  | 0 (0%) |  |  |
| Seroma |  | 2 (6.1%) | 3 (9.1%) |
| Seroma + Local Infection |  | 1 (3.0%) | 0 (0%) |

<sup>1</sup> p-value for the ANOVA test for continuous variables, the Kruskal-Wallis test for non-normal variables, and the Chi-Square test for categorical variables (or Fisher's exact test when appropriate). The letters followed by the p-values represent a statistically significant result for the Anova post-hoc test, with a: patent blue x patent blue + indocyanine green, b: patent blue x indocyanine green, c: patent blue + indocyanine green x indocyanine green. P-values in bold were statistically significant at the level $\alpha$ %.

**Fig 1.**
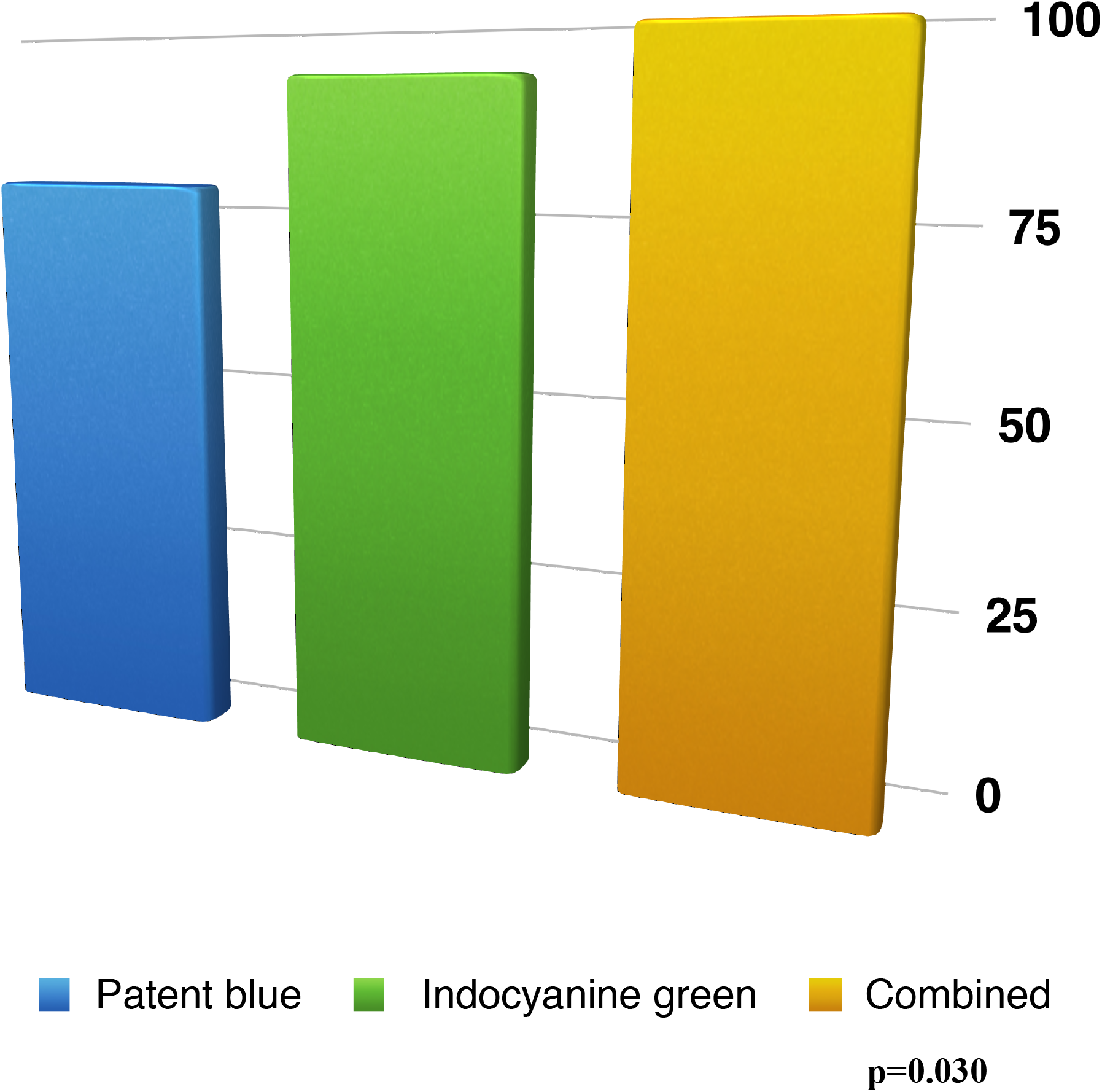
Sentinel lymph node identification rate in the patent blue, indocyanine green and combined arms.

The mean time of sentinel lymph node identification was 20.6 minutes among patients submitted to the traditional dye, 8.6 minutes in the indocyanine green arm and 10 minutes in the combination of the two methods (P<0.001). The mean surgery time was 69.4 minutes with patent blue, 55.1 minutes with indocyanine green and 69.4 minutes with the combination (P< 0.001) (Fig 2).

**Fig 2.**
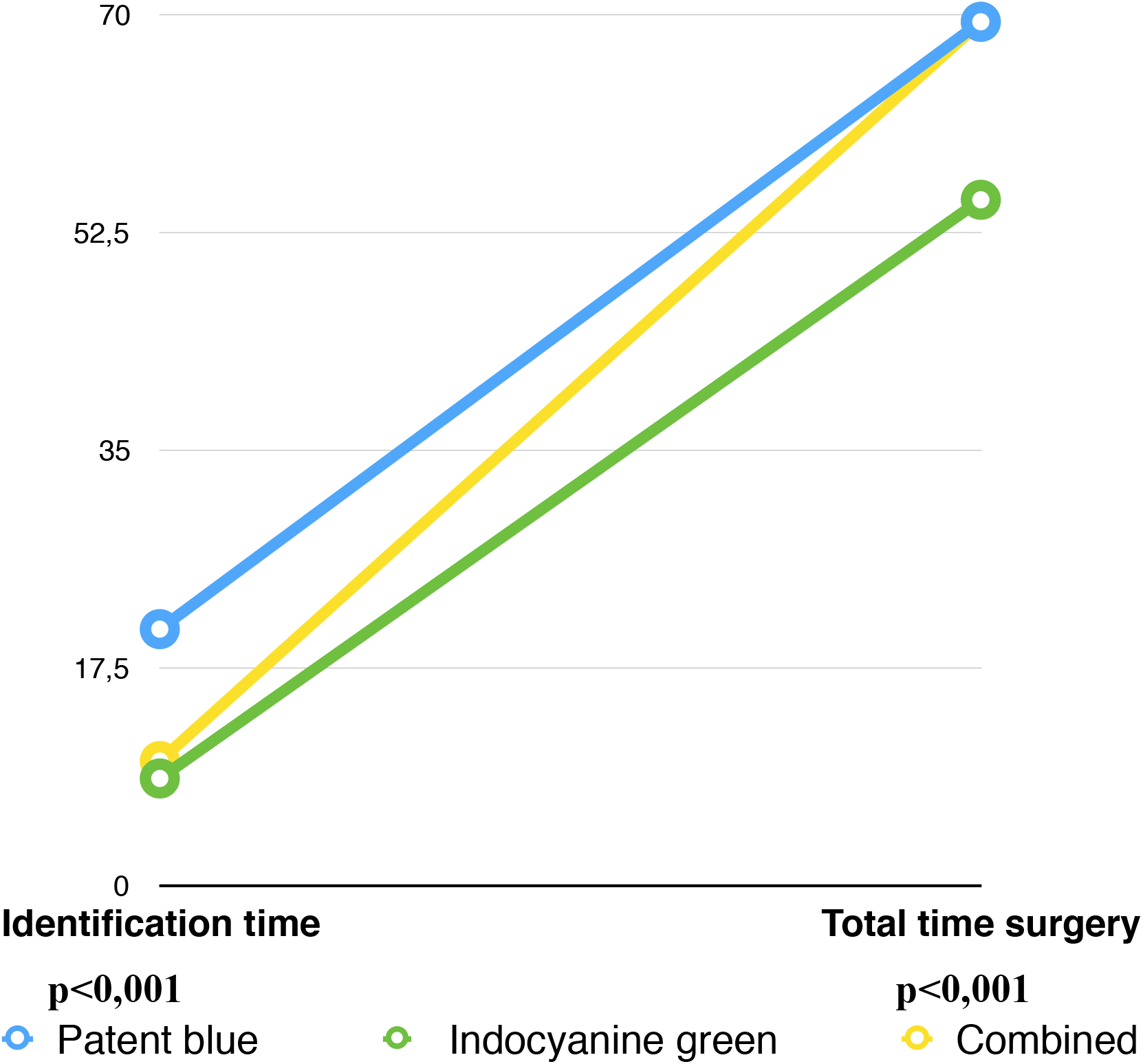
Rate of sentinel lymph node identification time and total surgery time in the patent blue, indocyanine green and combined technique arms.

The measurement of the breast incision was statistically significant only when it was single (P=0.020), with the smallest incision, 6.4 cm, in the indocyanine green arm. The measurement of the breast or axillary incision, when performed in association, that is, two incisions, did not reveal significant differences (Table 2).

The output of the Portovac drain in 7 days was also analyzed, with average measurements of 572.7 ml between patent blue, 448.8 ml in indocyanine green and 433.4 ml in the combined arm, thus not showing important differences (P=0.197) (Table 2).

Comparing the groups, the main postoperative adverse event was the dermal tattoo represented in 24 cases (72.7%) in the patent blue group (P=0.002). Among all patients who had a dermal tattoo, 71% corresponded to the patent blue arm (Fig 3).

**Fig 3.**
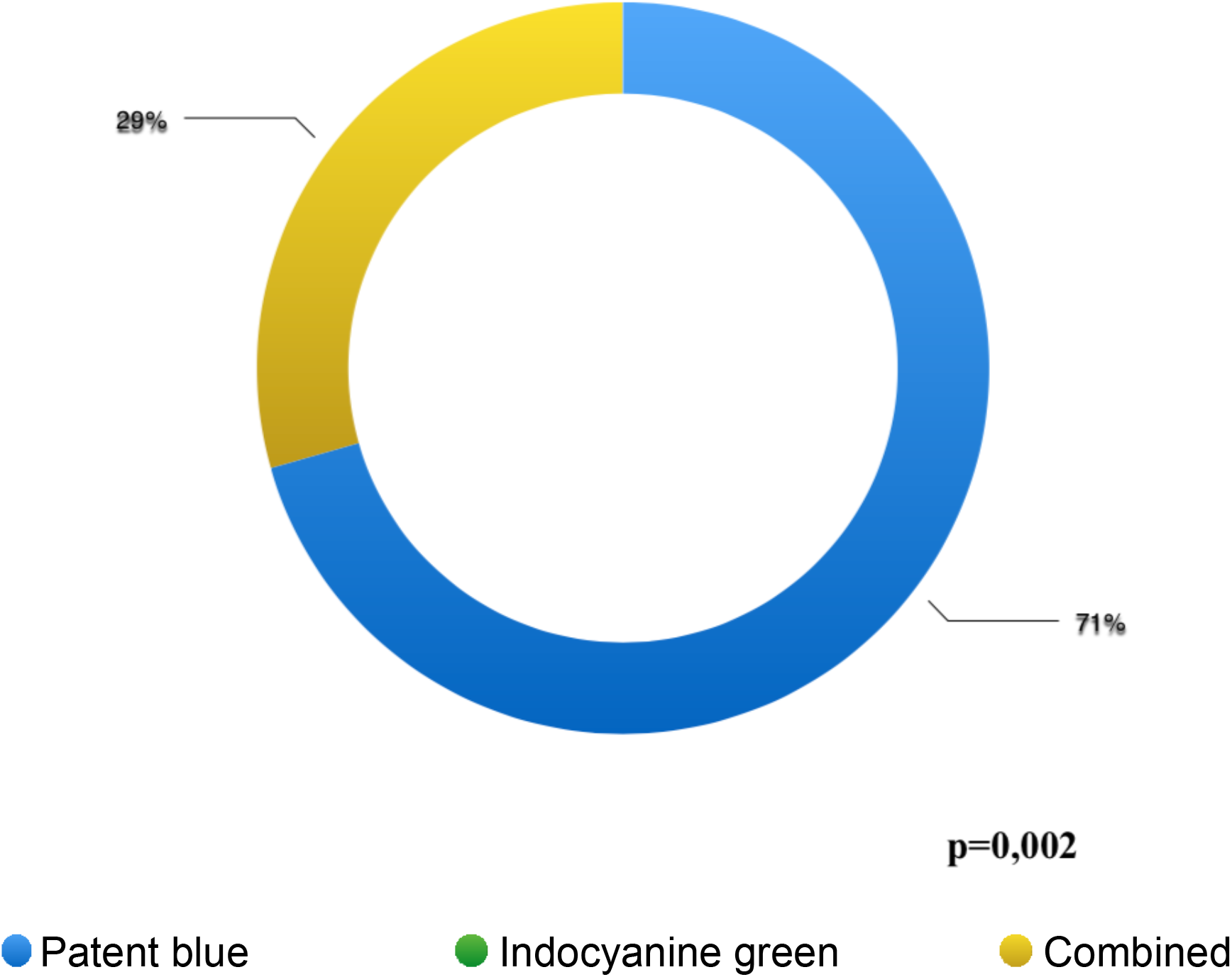
Dermal tattoo rate of the patent blue, indocyanine green and combined technique arms.

The amounts spent on surgeries performed with the patent blue dyes and the combined arm were not considered to be significantly different, while both were considered different from the expenses with surgeries performed with the green dye. This is noticeable because, on average, the amount spent on surgery with green dye was BRL 3,532, while with blue dye it was BRL 4,068 and with the combined, BRL 4,147 (Table 3). Although indocyanine green has a higher cost (BRL 150.00), the cost of hospitalization of patients submitted to this dye was lower than that of the patent blue dye p=0.013 (BRL 40.00) and the combined groups p= 0.003 (BRL 150.00 + BRL 40.00 = BRL 190.00). As for the comparison between the costs of the patent blue and combined arms, there was no statistically significant difference p=0.905 (Fig 4).

**Table 3.** Descriptive summary of values by dye.

| Dye | z | Average | Maximum | Sum | Standard Deviation | Groups* |
| --- | --- | --- | --- | --- | --- | --- |
| Blue | 1.930,83 | 4.068,34 | 7.170,70 | 134.255,2<br>0 | 870,91 | a |
| Green | 1.937,83 | 3.532,11 | 4.459,17 | 116.559,6<br>0 | 604,83 | b |
| Combined | 2.916,94 | 4.147,20 | 6.371,44 | 136.857,6<br>0 | zz | a |

\*According to the p-values of Tukey's multiple comparison test, $p=0.013$ for the blue-green comparison, $p=0.003$ for the combined green, and 0.905 for the combined blue.

**Fig 4.**
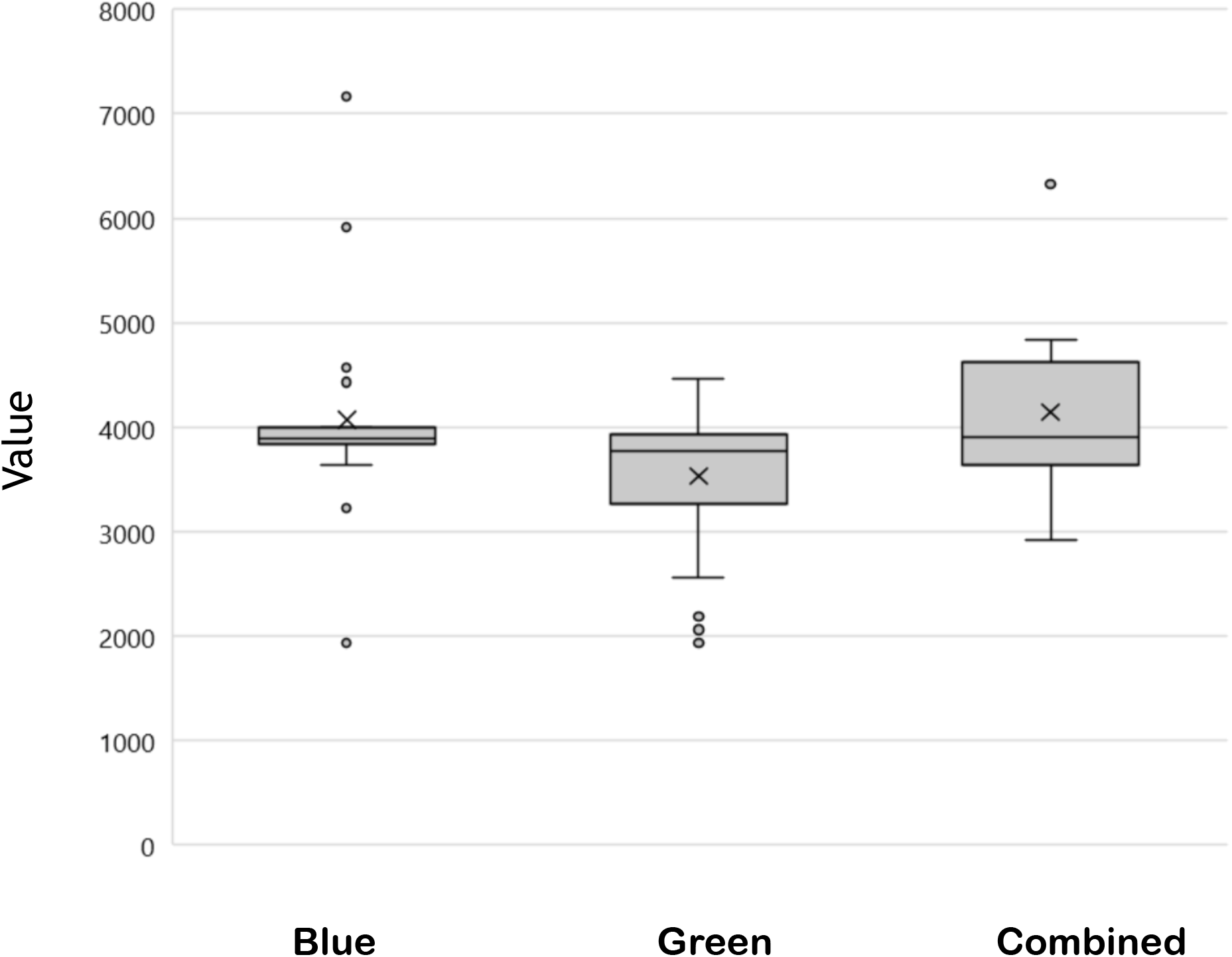
Comparison between the dye values in the three study arms, according to Tukey’s multiple comparison test.

## Discussion

Quadrantectomy was the main type of surgical treatment performed in patients in this study in all arms (81.8% in patent blue and combined; 90.9% in indocyanine green). Access to adjuvant radiotherapy provides this good rate of conservative surgery and reiterates the global trend of performing mastectomy only when precisely indicated and necessary^21^. Some patients who underwent mastectomy did not want immediate reconstruction (10.1% adding the 3 groups), even after the offer, explanation and encouragement. Our service has a team of reconstructive surgery and easy access to breast implants. However, the patient’s wishes were always respected. In the cases that underwent immediate breast reconstruction (5.1%), all underwent the expander technique. The different types of dyes for sentinel lymph node detection did not change the surgeons’ approach (type, location or number of incisions). The Mastology Team at Hospital de Esperança is composed of three breast surgeons, who were responsible for the procedures.

Primary surgery still has its role and was rightly proven in the study as the first indication in small (higher T1c rate in all groups) and luminal (luminal A + luminal B = 72.8%) tumors, as well as guided by the major global guidelines such as ASCO, ESMO and NC-CN ^14–16^. The majority presence of small tumors favored the prevalence of initial staging: anatomical staging IA in 53.5% and prognostic staging IA in 73.7%. This difference between anatomical staging and prognostic staging was due to the high participation of luminal tumors (stage IIA, which, because it is luminal, was reclassified to IA by the eighth edition of the AJCC)^20^. Most tumors were classified as invasive carcinomas of the non-special type, also in line with the literature ^22^.

The detection rate was surprisingly higher in patients who used fluorescence, reaching 100% when combined with patent blue. The aim of studying a new technique for sentinel lymph nodes was to provide real data so that cancer centers that do not have a nuclear medicine service may, in the future, have access to another option in addition to the renowned patent blue, which is still used in most hospitals, especially among users of the Unified Health System. Detection rates with indocyanine green are also high among major publications. Kitai et al^9^, a pioneer in its use, obtained a rate of 94% in Japan in 2004 (18 patients). Hirche et al^18^ reached 97.7% in Germany in 2010 (n=43) and Sorrentino et al^17^ 92.7% in Italy in 2018 (n=70).

The use of the combined technique, indocyanine green + patent blue, has already been tested by Shen et al^23^ in China and published in 2018 with a detection rate of 99.2%. A factor that could impact the detection rate would be the difference between the patients’ BMI or age; however, the distribution was homogeneous between the groups (BMI P=0.875, age P=0.220), with overweight (BMI between 25-29,9) and post-menopausal patients prevailing.

The dose of patent blue application is already consolidated in clinical practice, with the entire dye ampoule being usually infiltrated (5mg/2ml). However, even the main study, published by Giuliano et al, which described the use of the technique in the breast oncology scenario, showed divergence between the doses received by the patients: the first 20 patients received doses of 0.5 to 10 ml. From patient 21 to patient 172, the standard dose of the established dye was between 3 to 5 ml. This evidences the difficulty of determining the ideal dose of a new technique recently incorporated by the surgeon^12^.

The dose of ICG application and the camera used for detection is still heterogeneous in the literature, but the infiltration has always been described in the periareolar region. The pioneering Japanese study in the procedure used a dose of 25mg/5ml of the Diagnogreen® 0.5% brand (Daiichi Pharmatical, Tokyo Japan), being infiltrated soon after anesthesia and performing a two-minute breast massage, detected with the device from Hamamatsu Photonics® (Japan) ^9^. The German study applied the ICG Solution at a dose of 11mg, with massage for 5 to 15 minutes, with the IC-View®, Pulsion Medical Systems (Munich, Germany)^18^. The Italian study, in turn, drastically reduced the dose to 1.5mg of the ICG-Pulsion® dye (Pulsion Medical Systems (Munich, Germany)) with the D-Light P® 20cm camera coupled with the image1 S Camera Platform ® (Karl-Storz®, Germany), which was created for laparoscopy, but can also be used for open surgery ^17^; no details are described if the massage was performed. This material used in the Italian study is similar to this research and shares the characteristic of using a single system for different types of surgeries. The Chinese study, which evaluated the three arms similar to this research, used a dose of 6.5mg with the Photodinamic eye® camera (Hamamatsu Photonics, Japan), with a 10-minute massage ^23^. We emphasize here the difficulty in determining the exact time of the massage, which is described in a heterogeneous way by the literature, even in the scenario of the classic patent blue: Giuliano et al performed an interval of 1 to 20 minutes between the first 20 cases. From the twenty-first patient onwards, the standard time was defined as five minutes, which is currently followed by most oncology services^4,12^.

This study was a pioneer in comparing the time of sentinel lymph node identification among the dyes used. The advent of fluorescence also had the benefit of allowing less time to visualize the first lymph node of the axillary drainage and its excision (12-minute reduction compared to patent blue). Therefore, the mean time of the surgical procedure was also reduced in relation to other dyes (14.3-minute reduction). Surgery time was similar between the patent blue and combined arms. The main benefit in reducing sentinel lymph node identification time and surgery time is the shorter time the patient is exposed to general anesthesia and mechanical ventilation, which may reduce the risk of intraoperative and postoperative complications, in addition to the lower cost of anesthetic drugs.

Despite the research, the team of surgeons focused on not changing their surgical indications, as well as maintaining the same type, quantity and topography of the incisions. Classically, the sentinel lymph node technique is performed through an additional axillary incision ^23^. However, currently breast surgeons aim for better cosmetic results whenever possible, and some patients receive only a dermal scar for both breast cancer surgery and axillary surgery ^24^. Following this trend, more than half of the cases received only one breast incision. Regarding the number of incisions, the three groups were homogeneous (p=0.849). The single incision had a smaller measurement in the indocyanine green group, 3.4 cm less than with the use of patent blue alone and 1.9 cm less than the combined technique. This fact, which was statistically significant (p=0.020), may have an impact on a lesser breast mutilation aspect in the postoperative period and better cosmesis.

We realized that the ICG is ultrasensitive and migrates through the lymphatic pathway extremely quickly. However, in some cases we observed that the dye leaked into adjuvant tissues to the lymph nodes. Thus, we believe that a lower dose can be sufficient and also efficient. We aim to strictly maintain the massage dose and time proposed in the research project, in order to guarantee research homogeneity. New studies may determine a lower and ideal dose, even encouraging the pharmaceutical industry to market vials with the ideal presentation for the lymph node scenario.

Among the 66 patients who used indocyanine green (33 in the isolated arm and 33 in the combined arm), none presented allergy, which suggests that this atopy is extremely rare. However, for safety reasons, the anamnesis had already investigated the allergy to iodides and the anesthesiologist was prepared for a possible reversal of the atopic condition, if necessary. The fact that the drain output did not show a difference in the groups suggests that the type of dye does not interfere with the production of seroma in the first postoperative week. The dermal tattoo caused by patent blue does not interfere with the oncological treatment, but it generates visual discomfort of the breast skin for the patient and her surgeon, which is the main statistically significant complication: more than two thirds of the cases of dermal tattoo were caused by patent blue (71%, p=0.002). The use of indocyanine green generated a higher rate of late seroma in relation to the other groups. However, the incidence was low, although the percentage was higher; in gross numbers, it presented only one case more than the combined arm and two more cases than the patent blue arm.

Our device is still at the forefront of this technology. However, new technologies have been incorporated for commercialization and today there is already a more modern device, also from Karl-Storz®, called IMAGE1 S™ RUBINA™, which already enables 3D and 4K images ^25^. We know that not all hospitals have this fluorescence equipment, which is still expensive. However, with the advent of robotics, some of these devices already include fluorescence technology and their use could be offered to breast surgeons ^26^. In addition to consolidating national data on the use of the ICG, we aim to disseminate it widely to other Mastology and Oncological Surgery services so that more patients and surgeons can also benefit from learning this technique. For hospitals that have not yet had access to the high technology of fluorescence, an option in the near future is the commercialization of fluorescence devices of national manufacture, which are still prototypes and have been tested in head and neck surgery at Hospital de Amor with promising results ^27^.

Indocyanine green already has a scientific description, although still rudimentary, of therapeutic effect, such as in the treatment of acne and even in metastatic nodules ^10,28^. We hope to publish, in the future, a graph of disease-free survival and overall survival from the 60-month follow-up of patients participating in this research, in order to study this possible mechanism. So far, we have revealed the data that one patient had local recurrence, seven had distant metastases, and none of these cases belonged to the indocyanine green arm. Shen et al already published there is no statistically significant difference in disease-free survival at 50 months of follow-up in patients submitted to patent blue (n=149) and patent blue + ICG (n= 374), p= 0.322 ^23^.

The monetary value of hospitalization for each patient in the study was performed in order to confirm that the cost of a more modern dye would not have a financial impact on the institution.

One of the reasons that explains the unusual finding in the comparison with patent blue is the fact that, by chance, breast reconstruction was not performed in patients who used indocyanine green. The cost of material such as the breast implant and the fees for breast reconstruction contribute to the increase in the overall cost of hospitalization. It is important to emphasize that the cost of the fluorescence equipment is high; however, this equipment already belonged to the structure of the surgical center of Hospital de Esperança before the research was started and we had no additional costs for using this technology.

## Conclusions

The sentinel lymph node detection rate by fluorescence with the use of indocyanine green was 93.9%, being considered effective. The comparison of the sentinel lymph node detection rate between the use of patent blue, indocyanine green and patent blue + indocyanine green (combined) revealed statistically significant differences (p=0.030), with the combined method being the most effective with a rate of 100%. The higher cost of the indocyanine green dye did not impact the increase in the overall cost of hospitalization.

## Data Availability

All data produced in the present study are available upon reasonable request to the authors.

## Acknowledgments

The first author (RSS) was supported as a Doctoral Scholar by the National Council for Scientific and Technological Development (CNPq). This study received no other funding.

## Supporting information

**S1 Table. Characteristics of patients and tumors in the three study arms (patent blue, indocyanine green and combined)**.

**S2 Table. Comparison of the characteristics of patients and tumors in the three study arms (patent blue, indocyanine green and combined)**.

