## Supplementary material for "Evaluation of the efficacy of using indocyanine green associated with fluorescence in sentinel lymph node biopsy": S1 table

### S1 - Characteristics of patients and tumors in the three study arms (patent blue, indocyanine green and combined).

| Characteristic | n = 99 (%) |
|---|---|
| **Age** | |
| Mean ± standard deviation | 58.4 ± 13.3 |
| < 50 years | 29 (29.3%) |
| ≥ 50 years | 70 (70.7%) |
| **BMI** | |
| 18.5 – 24.9 | 27 (27.3%) |
| 25 – 29.9 | 44 (44.4%) |
| ≥ 30 | 28 (28.3%) |
| **Region** | |
| Right inferior lateral quadrant | 2 (2.0%) |
| Left inferior lateral quadrant | 5 (5.0%) |
| Left inferior medial quadrant | 1 (1.0%) |
| Right superior lateral quadrant | 21 (21.2%) |
| Left superior lateral quadrant | 14 (14.1%) |
| Right superior medial quadrant | 7 (7.1%) |
| Left superior medial quadrant | 6 (6.1%) |
| Right retroareolar region | 4 (4.0%) |
| Left retroareolar region | 8 (8.1%) |
| Union of the lateral quadrants of the right breast | 6 (6.1%) |
| Union of the upper quadrants of the right breast | 7 (7.1%) |
| Union of the upper quadrants of the left breast | 7 (7.1%) |
| Union of the lower quadrants of the right breast | 1 (1.0%) |
| **Characteristic** | **n = 99 (%)** |
| Union of the lower quadrants of the left breast | 2 (2.0%) |

| | |
|---|---|
| Union of the lateral quadrants of the left breast | 8 (8.08%) |
| **Type of surgery** | |
| Breast-conserving surgery (quadrantectomy) with sentinel lymph node biopsy | 84 (84.8%) |
| Mastectomy with sentinel lymph node biopsy with breast reconstruction (expander) | 5 (5.1%) |
| Mastectomy with sentinel lymph node biopsy without reconstruction | 10 (10.1%) |
| **Anatomical Staging** | |
| IA | 53 (53.5%) |
| IIA | 27 (27.3%) |
| IIIA | 4 (4.0%) |
| IIB | 12 (12.1%) |
| IIIC | 3 (3.0%) |
| **Prognostic Staging** | |
| IA | 73 (73.7%) |
| IIA | 10 (10.1%) |
| IB | 12 (12.1%) |
| IIB | 2 (2.0%) |
| IIIB | 2 (2.0%) |
| **Tumor size** | |
| 6-10mm (T1b) | 13 (13.1%) |
| 11-20mm (T1c) | 53 (53.5%) |
| 21-50mm (T2) | 33 (33.3%) |
| **Histology** | |
| Invasive carcinoma of no special type | 92 (92.9%) |
| Invasive lobular carcinoma | 4 (4.0%) |
| Invasive medullary carcinoma | 1 (1.0%) |

| **Characteristic** | **n = 99 (%)** |
|---|---|
| Invasive papillary carcinoma | 2 (2.0%) |

| | |
|---|---|
| **Grade** | |
| 1 | 5 (5.1%) |
| 2 | 64 (64.6%) |
| 3 | 30 (30.3%) |
| **Immunohistochemistry** | |
| HER2+ | 7 (7.1%) |
| Luminal A | 37 (37.4%) |
| Luminal B | 35 (35.4%) |
| Luminal HER | 12 (12.1%) |
| Triple Negative | 8 (8.1%) |
