## Supplementary material for "Evaluation of the efficacy of using indocyanine green associated with fluorescence in sentinel lymph node biopsy": S2 table

### S2 - Comparison of the characteristics of patients and tumors in the three study arms (patent blue, indocyanine green and combined).

| Characteristic | Patent blue (n=33) | Patent blue + indocyanine green (n=33) | Indocyanine green (n=33) | P-value[1] |
|---|---|---|---|---|
| **Age** | | | | |
| Mean ± standard deviation | 58.7 ± 13.9 | 59.6 ± 13.3 | 57.0 ± 13.0 | 0.717 |
| < 50 years | 11 (33.3%) | 6 (18.2%) | 12 (36.4%) | |
| ≥ 50 years | 22 (66.7%) | 27 (81.8%) | 21 (63.6%) | 0.220 |
| **BMI** | | | | |
| 18.5 – 24.9 | 10 (30.3%) | 7 (21.2%) | 10 (30.3%) | |
| 25 – 29.9 | 15 (45.5%) | 16 (48.5%) | 13 (39.4%) | 0.875 |
| ≥ 30 | 8 (24.2%) | 10 (30.3%) | 10 (30.3%) | |
| **Region** | | | | |
| Right inferior lateral quadrant | 0 (0%) | 1 (3%) | 1 (3%) | |
| Left inferior lateral quadrant | 2 (6.1%) | 2 (6.1%) | 1 (3%) | |
| Left inferior medial quadrant | 0 (0%) | 0 (0%) | 1 (3%) | |
| Right superior lateral quadrant | 11 (33.3%) | 5 (15.2%) | 5 (15.2%) | |
| Left superior lateral quadrant | 4 (12.1%) | 3 (9.1%) | 7 (21.2%) | |
| Right superior medial quadrant | 2 (6.1%) | 4 (12.1%) | 1 (3%) | |
| Left superomedial quadrant | 1 (3%) | 1 (3%) | 4 (12.1%) | |
| | 2 (6.1%) | 2 (6.1%) | 0 (0%) | |

| | | | | |
|---|---|---|---|---|
| Right retroareolar region | | | | **0.026** |
| Left retroareolar region | 4 (12.1%) | 2 (6.1%) | 2 (6.1%) | |
| Union of the lateral quadrants of the right breast | 1 (3%) | 3 (9.1%) | 2 (6.1%) | |
| Union of the upper quadrants of the right breast | 2 (6.1%) | 1 (3%) | 4 (12.1%) | |
| Union of the upper quadrants of the left breast | 2 (6.1%) | 5 (15.2%) | 0 (0%) | |
| Union of the lower quadrants of the right breast | 0 (0%) | 1 (3%) | 0 (0%) | |
| Union of the lower quadrants of the left breast | 2 (6.1%) | 0 (0%) | 0 (0%) | |
| Union of the lateral quadrants of the left breast | 0 (0%) | 3 (9.1%) | 5 (15.2%) | |
| **Type of surgery** | | | | |
| Breast-conserving surgery (quadrantectomy) with sentinel lymph node biopsy | 27 (81.8%) | 27 (81.8%) | 30 (90.9%) | 0.555 |
| Mastectomy with sentinel lymph node biopsy with breast reconstruction (expander) | 2 (6.1%) | 3 (9.1%) | 0 (0%) | |
| Mastectomy with sentinel lymph node biopsy without reconstruction | 4 (12.1%) | 3 (9.1%) | 3 (9.1%) | |
| **Anatomical Staging** | | | | |
| I A | 17 (51.5%) | 14 (42.4%) | 22 (66.7%) | |
| I I A | 7 (21.2%) | 12 (36.4%) | 8 (24.2%) | |
| IIIA | 0 (0%) | 3 (9.1%) | 1 (3.0%) | 0.063 |
| I I B | 8 (24.2%) | 2 (6.1%) | 2 (6.1%) | |
| IIIC | 1 (3.0%) | 2 (6.1%) | 0 (0%) | |
| **Prognostic Staging** | | | | |
| I A | 19 (57.6%) | 27 (81.8%) | 27 (81.8%) | |
| IIA | 5 (15.2%) | 3 (9.1%) | 2 (6.1%) | |
| IB | 8 (24.2%) | 1 (3.0%) | 3 (9.1%) | 0.090 |

| | | | | |
|---|---|---|---|---|
| IB | 8 (24.2%) | 1 (3.0%) | 3 (9.1%) | 0.090 |
| IIB | 0 (0%) | 1 (3.0%) | 1 (3.0%) | |
| IIIB | 1 (3.0%) | 1 (3.0%) | 0 (0%) | |
| **Tumor size** | | | | |
| 6-10mm (T1b) | 1 (3%) | 4 (12.1%) | 8 (24.2%) | |
| 11-20mm (T1c) | 18 (54.5%) | 16 (48.5%) | 19 (57.6%) | **0.046** |
| 21-50mm (T2) | 14 (42.4%) | 13 (39.4%) | 6 (18.2%) | |
| **Histology** | | | | |
| Invasive carcinoma of no special type | 31 (93.9%) | 29 (87.9%) | 32 (97.0%) | |
| Invasive lobular carcinoma | 1 (3.0%) | 3 (9.1%) | 0 (0%) | |
| | | | | 0.346 |
| Invasive medullary carcinoma | 0 (0%) | 0 (0%) | 1 (3.0%) | |
| Invasive papillary carcinoma | 1 (3.0%) | 1 (3.0%) | 0 (0%) | |
| **Grade** | | | | |
| 1 | 1 (3.0%) | 1 (3.0%) | 3 (9.1%) | |
| 2 | 22 (66.7%) | 22 (66.7%) | 20 (60.6%) | 0.786 |
| 3 | 10 (30.3%) | 10 (30.3%) | 10 (30.3%) | |
| **Immunohistochemistry** | | | | |
| HER2+ | 1 (3.0%) | 4 (12.1%) | 2 (6.1%) | |
| Luminal A | 13 (39.4%) | 11 (33.3%) | 13 (39.4%) | |
| Luminal B | 15 (45.5%) | 12 (36.4%) | 8 (24.2%) | 0.377 |
| Luminal HER | 2 (6.1%) | 5 (15.1%) | 5 (15.1%) | |
| Triple Negative | 2 (6.1%) | 1 (3.0%) | 5 (15.1%) | |

[1] p-value for the ANOVA test for continuous variables, the Kruskal-Walis test for non-normal variables, and the Chi-Square test for categorical variables (or Fisher's exact test when appropriate). The letters followed by the p-values represent a statistically significant result for the Anova post-hoc test, with a: patent blue x patent blue + indocyanine green, b: patent blue x indocyanine green, c: patent blue + indocyanine green x indocyanine green. P-values in bold were statistically significant at the level $\alpha\%$.
